# Exposome approaches to assessing the association between urban land use environment and depressive symptoms in young adulthood: a FinnTwin12 cohort study

**DOI:** 10.1101/2023.03.27.23287783

**Authors:** Zhiyang Wang, Alyce M.Whipp, Marja Heinonen-Guzejev, Jordi Júlvez, Jaakko Kaprio

## Abstract

**Background:** Depressive symptoms lead to a serious public health burden and are considerably affected by the environment. Land use, describing the urban living environment, has an impact on mental health, but complex relationship assessment is rare.

**Objectives:** We aimed to examine the complicated association between urban land use and depressive symptoms among young adults with differential land use environments, by applying multiple models, as an exposome study.

**Methods:** We included 1804 individual twins from the FinnTwin12 cohort, living in urban areas in 2012. There were 8 types of land use exposures in 3 buffer radii. The depressive symptoms were assessed through General Behavior Inventory (GBI) in young adulthood (mean age: 24.1). First, K-means clustering was performed to distinguish participants with differential land use environments. Then, linear elastic net penalized regression and eXtreme Gradient Boosting (XGBoost) were used to reduce dimensions or prioritize for importance and examine the linear and nonlinear relationships.

**Results:** Two clusters were identified with notable differences in the percentage of high-density residential, low-density residential, and natural land use. One is more typical of city centers, and another of suburban areas. A heterogeneous pattern in results was detected from the linear elastic net penalized regression model among the overall sample and the two separated clusters. Agricultural residential land use in a 100 m buffer contributed to GBI most (coefficient: 0.097) in the “suburban” cluster among 11 selected exposures. In the “city center” cluster, none of the land use exposures was associated with GBI. From the XGBoost models, we observed that ranks of the importance of land use exposures on GBI and their nonlinear relationships are also heterogeneous in the two clusters.

**Discussion:** As a hypothesis-generating study, we found heterogeneous linear and nonlinear relationships between urban land use environment and depressive symptoms under different contexts in pluralistic exposome analyses.

## Introduction

Depressive symptoms are very common and reflect a chronic, complex, and multifactorial mental health condition. The burden of depressive symptoms is growing especially among younger people. A national survey in the U.S. showed that there is a large rise in the incidence of major depressive episodes among young adults.^1^ A recent survey in Spain suggested that 23.6% of college students experienced depressive symptoms^2^, while the prevalence was 28.4% among Chinese university students by a systematic review.^3^ The COVID-19 pandemic induced a negative mental health impact and increased the prevalence of depressive symptoms among young adults.^4,5^ Moreover, depressive symptoms have been associated with a higher odd of risk behavior such as substance use and self-harm, which resulted in further psychological and physical health problems.^6^ Several twin studies across countries have identified the major role of environmental influences on mental health, including depressive symptoms among young adults, inspiring etiological consideration of people’s various environments.^7,8^

Land use involves the transformation of undeveloped areas into a sound and vital residential and living environment. Urban planners consider multiple concepts such as suitability, competitiveness, need diversity, or resource scarcity to evaluate land use.^9^ Furthermore, in the “One Earth” perspective, land use is closely connected to biodiversity and agriculture, which are reciprocally related to people.^10^ Thereby, advancing liveable initiatives and shaping diverse land use is able to promote healthy lifestyles, urban amenities, and nature conservation, ultimately leading to a better Earth.^11,12^ Some studies have addressed the relationship between land use and mental health/status. Miles et al. assessed the association between land use diversity, via Herfindahl–Hirschman Index, and depressive symptoms among Miami residents in US., but there was no salient result.^13^ An Italian study also found that land use mix, calculated via the Shannon diversity index, was not significantly associated with prescriptions of antidepressants.^14^ Nevertheless, land use mix, measured by the entropy model, was demonstrated to be correlated with life satisfaction at residences and workplaces in Beijing, China.^15^ Existing indices have some limitations, such as insensitiveness to capture the land use interaction.^16^ Inconsistent evidence reflects the complexity of the land use effect, which demands further sophisticated analysis.

The urban exposome describes the totality of environmental exposure that people experience on a daily basis in cities as an important component in the external exposome, and over 75% of the European population lives in urban areas. As a part of the urban exposome, studies on land use also encounter difficulties such as high-dimensionailty and pleiotropy.^17^ Instead of conventional regression models with a single index, interpretable and robust multi-exposure models are recommended. Ohanyan and colleagues have built some machine learning models, illustrated their characteristics, and applied them to a study on the urban exposome and type-2 diabetes.^18,19^ However, this type of research is rarely used on mental health. To fulfill the current research gap, we conducted this exposome study with three objectives: a) to cluster participants who shared a similar pattern of urban land use; b) to assess both the linear and non-linear relationships between urban land use and depressive symptoms in young adulthood; and c) to observe the possible differences in these relationships between clusters.

## Methods

### Study participants

The participants were from the FinnTwin12 cohort, which is a population-based prospective cohort among all Finnish twins born between 1983 and 1987, and their parents. At baseline, 5522 twins were invited and 5184 twins replied to our questionnaire (age 11–12, wave one), and they compose the overall cohort. All twins were invited to participate in the first follow-up survey with 92% retention at age 14 (wave two). Moreover, at age 14, 1035 families were invited to take part in an intensive substudy with psychiatric interviews, some biological samples, and additional questionnaires, and of 1854 twins participated in these interviews. They were also invited to a second intensive survey as young adults, with a participation rate of 73% (n=1347 individual twins), and completed the detailed young adulthood questionnaires and interviews (part of wave four). In addition, all of the twins in the overall cohort completed general age 17 questionnaires (wave three) and twins from the non-intensive study completed young adult questionnaires (wave four) with 75% and 66% retention, respectively. In this study, we included twins who participated in wave four. An updated review of this cohort was published recently.^20^

### Measures

#### Depressive symptoms

In this study, the short-version General Behavior Inventory (GBI) was used to evaluate depressive symptoms among twins in young adulthood.^21^ It is a self-reported inventory designed to identify mood-related behaviors, which is composed of 10 questions with a 4-point Likert scale from 0 (never) to 3 (very often) to query the occurrence of depressive symptoms.^22^ The total score ranges from 0 to 30, and a higher score implies more depressive symptoms occurred. To validate the GBI, we compared it to a Diagnostic and Statistical Manual of Mental Disorders-IV diagnosis of major depressive disorder (MDD) assessed by the Semi-Structured Assessment for the Genetics of Alcoholis (SSAGA) m interview from the intensive study.^23^ In a logistic regression model, the GBI score in young adulthood strongly predicted MDD, with the area under the receiver operating characteristic curve (AUC) of 0.8328 (among twins included in this study’s analysis).

#### Land use

The EUREF-FIN geocodes of twins from birth to 2021 were derived from the Digital and Population Data Services Agency, Finland. We used geocodes in 2012 to merge the land use exposures to twins, derived from Urban Altas 2012. Urban Altas is a part of land monitoring services to provide reliable, inter-comparable, high-resolution land use maps in the European Union and European Free Trade Association countries from 2006 to 2016, which covered nearly 700 larger functional urban areas in 2012.^24^ Land use exposures included the percentage of 8 types of land use (high-density residential, low-density residential, industrial and commercial, infrastructure, urban green, agricultural, natural, and water) within an area of 100, 300, and 500 m radius buffer zones for each geocode in urban Finland (totally 24 exposures).

Additionally, we also calculated the land use mix index within different buffers, which described the diversity of land uses through Shannon’s Evenness Index. The equation is defined as follows:^25^

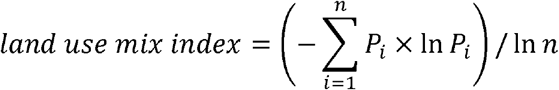

*P*_i_ is the percentage of each type of land use in zone *i*; *n* is the number of land use types. It ranges from 0 to 1, and a higher value indicates a more balanced distribution of land between the different types of land use.

#### Covariates

Seven covariates were defined *a priori*: sex (male, female), zygosity (monozygotic (MZ), dizygotic (DZ), unknown), parental education (limited, intermediate, high), smoking (never, former, occasional, current), work status (full-time, part-time, irregular, not working), secondary level school (vocational, senior high school, none), and age. The latter four variables came from the young adulthood survey (mean age at response based on the difference of date of response and data of birth: 24.07 years). Parental education was based on maternal and paternal reports, while zygosity was based on DNA polymorphisms and/or a validated zygosity questionnaire.^26^

### Analysis

#### Preparation and description

We only included the twins who have available land use exposures in 2012 in urban areas (as defined above), indicating that they lived in the urban areas in Finland, and provided GBI assessment in young adulthood, in order to have a larger sample size and have the two measurements be as close as possible on the time scale. A total of 1804 individual twins (589 twin pairs and 626 individual twins) were included and the mean age in providing GBI assessment was 24.07 years (around 2007-2011). Due to the skewness of the GBI score, we add one to the GBI score and log-transformed it for the following analysis. A correlation matrix was drawn between land use exposures. Then, we proposed a two-stage exposome approach to assess the relationship between land use exposures and depressive symptoms.

#### Stage 1: unsupervised clustering

To group twin individuals who have similar land use in an exploratory way, we used unsupervised K-means clustering. The K-means clustering method employs a non-hierarchical partitional algorithm. It calculated the total within-cluster variation as the sum of the squared Euclidean distance between each sample and the corresponding K-number random-assigned centroid in each cluster (*k*). *X*_*ik*_ is the i^th^ observation belonging to cluster (*k* = 1, 2, ….,, K) and *n*_*k*_ is the number of observations in cluster *k*. The overall within-cluster variation is defined as follows:^27^

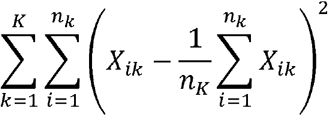

The process will stop when a convergence criterion is met (smallest overall within-cluster variation).^27^ It is one of the simplest and fastest clustering methods, and is also able to handle outliers or inappropriate variables.^28,29^ Only the 24 land use exposures were included in the clustering algorithm. We used the Silhouette method to estimate the optimal number of pre-specified cluster^30^, and two clusters were identified (Supplemental Figure 1). The R package “Factoextra” was used.^29^

#### Stage 2: Exposome pluralistic analysis

We split the twin participants into training and testing subsets. In full twin pairs, we performed a 1:1 random split within the pair. The remaining individual twins all went to the training subset. The training sample size was 1215 and the testing sample size was 589, and the size in each cluster varied (Supplemental Table 1). By the splitting process, we do not need to consider the statistical effect of complex sampling cluster effects by twin pair status as all individuals in both samples are unrelated. We chose two types of models and adjusted covariates to evaluate the risk estimation of 24 land use exposures (*j*).

First, the linear elastic net penalized regression model was applied for feature selection, which uses a hybrid of the lasso and ridge penalized methods, to fit the generalized linear model.^31^ This model considered multicollinearity by removing any degeneracies and outlying behavior and assessed the linear relationship.^32^ A typical linear regression model based on N participants with the combined penalized term is defined as follows (cited from Fridman at el.^32^):

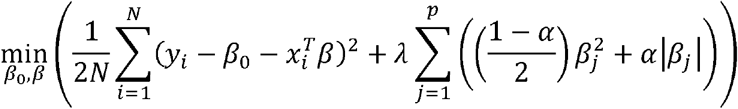

*y*_*i*_ is the dependent response and *x*_*i*_ is the independent factor at observation *i*. λ is a positive regularization parameter. *β*_0_ and *β* are scalar and p-vector coefficients, respectively. We set the *α* ranging from 0.1 to 1.0, as a tuning parameter, for the penalty. We fixed the covariates in the models as unpenalized variables to fully adjust them. The final models were selected by 10-fold cross-validation to determine the optimal degree of penalization.^31^ Stata package “elasticnet” was used.

Further, to assess the non-linearity relationship, the supervised machine learning model –– eXtreme Gradient Boosting (XGBoost) was used. It is a tree-based gradient boosting technique, utilizing the weights of trees, which is good in predicting and less susceptible to overfitting.^33,34^ The objective function of XGBoost starts with two parts: a loss function and a regularization term, and we aim to obtain the optimal output value (*O*_*value*_) to minimize the function, defined as follows:

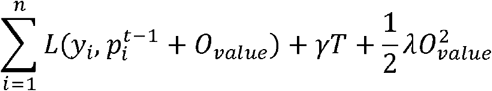

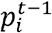 is the previous prediciton of tree *t* at observation *i. T* is the number of leaf nodes in a tree, and*γ* and *λ* are the definable penalty factors to avoid overfitting. Then, we rewrite the loss function according to the 2^nd^ Taylor Approximation:

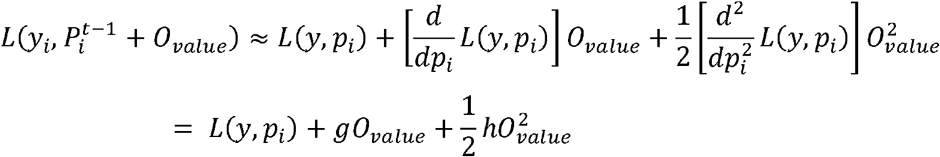

*L*(*y, p*_*i*_) is the loss function of the previous prediction, and its first and second derivative is labeled as *g* and *h*, respectively. The optimum output value could then be derived with *G* and *H* (sum of *g* and *h*) as:

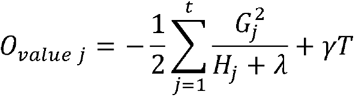

The detailed mathematical model and algorithm are described in previous literature.^35^ This model is able to characterize the interaction and nonlinearity.^18^ The tuning hyperparameters were calibrated by parallelizable Bayesian optimization based on 7 initialization evaluations and 30 epochs (50 epochs for Cluster 2), using the R package “ParBayesianOptimization”.^36,37^ We ran training XGboost models with 3000 rounds at first, then the optimal number of rounds (*n*) was selected by mean-squared error (MSE) as the following equation:

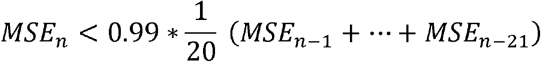

The Final XGBoost analysis was conducted with all hyperparameters using the R package “xgboost”.^33^ Covariates were included in the model. Finally, we used the Shapley (SHAP) value to interpret and visualize the results from the XGboost machine learning model with higher transparency by the R package “shapr”.^38,39^

Models were performed among overall participants and in two clusters. We used root-mean-squared error (RMSE) to measure model performance in the training and testing subsets, which is a weighted measure calculated between forecast and observed values.

#### Post-hoc analysis

We conducted a post-hoc linear regression between the land use mix index and log-transformed GBI score, which aims to compare with our novel findings. Covariates were adjusted for and the cluster effect of sampling based on families of twin pairs was controlled by the robust standard error. A p-value less than 0.05 is considered statistically significant and 95% confidence intervals (CI) are reported.

## Results

### K-means clustering and descriptive statistics

Figure 1 depicts the distribution of each land use category overall and in the two clusters. Cluster 2 had a higher percentage of high-density residential land use, while Cluster 1 had a higher percentage of low-density residential land use regardless of the buffer radii of the twins’ location. Supplemental Figure 2 shows the twins’ location in the greater Helsinki areas (as an example), and twins from Cluster 2 lived in more urbanized areas (often close to city or town centers), while twins from Cluster 1 were more suburban. Variable names and details are shown in Supplemental Table 2. We also calculated the simple ratios of means between the two clusters and found low-density residential, agricultural residential, and natural land use in a 100 m buffer have notably “relative” differences between the two clusters (ratio>10). According to the correlation matrix based on the training subset (Supplemental Figure 3), the same land use with different radii of the buffer zone is highly correlated. High-density and low-density residential land use are negatively correlated.

**Figure 1:**
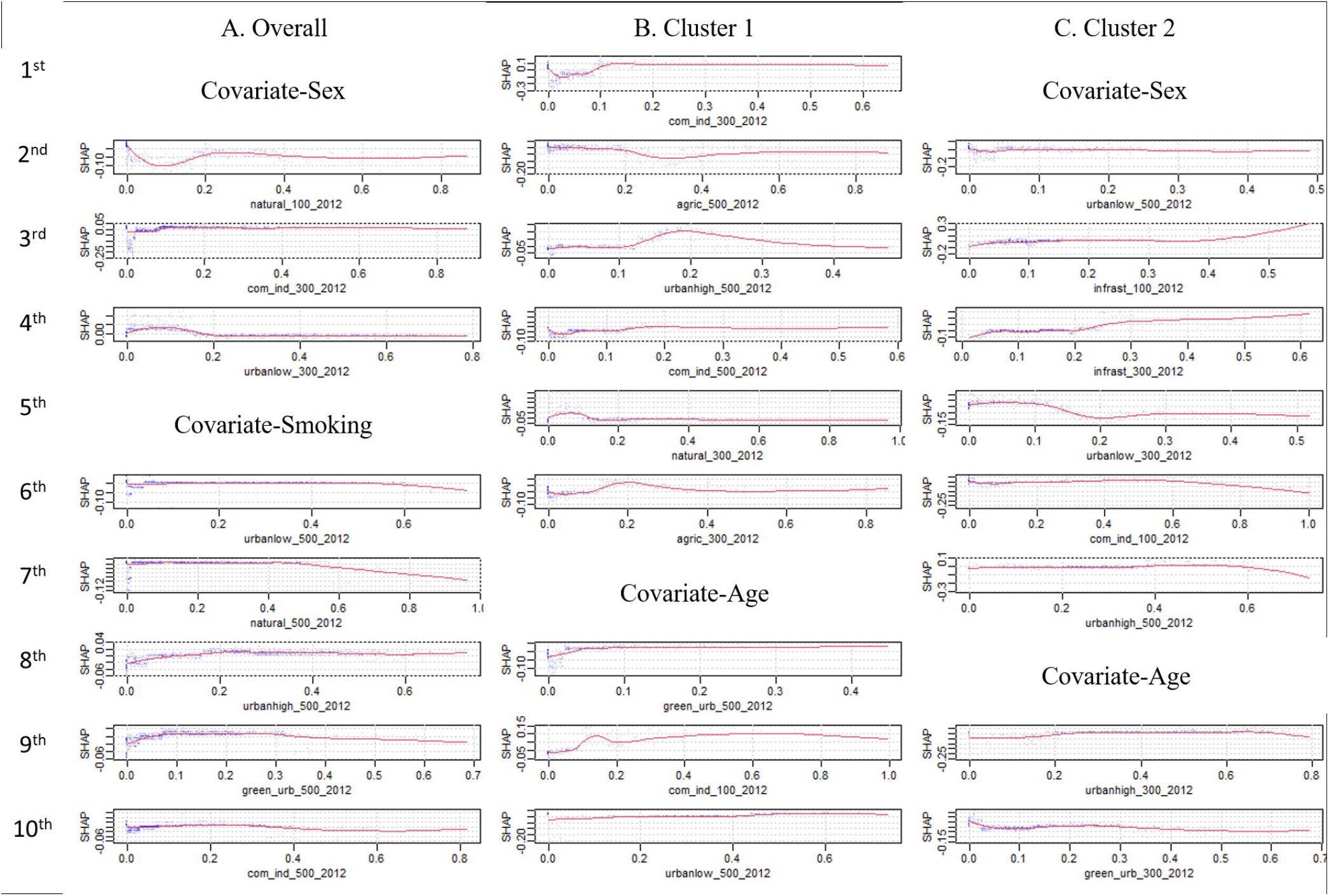
Histogram of the percentage of land use exposures among overall twins and in the two clusters

Table 1 shows the distribution of characteristics overall and in the two clusters. Overall, the majority of twins are female (58.7%), dizygotic (61.3%), and reported never smoking (55.1%) in the young adulthood questionnaire. Additionally, 48.8% and 47.7% of twins reported that they were in full-time work and had attended senior high school, respectively. The majority (51.1%) of twins’ parents had limited education levels (less than high-school). Unsupervised K-means clustering did not take into account these demographics covariates. We observed significant differences in smoking, working status, secondary level school, and parental education between the two clusters by Chi-squared test or univariable linear regression accounting for twin sampling. There were more twins who currently smorked, worked full time, and attended vocational schools in Cluster 1 (suburban) than in Cluster 2 (city center), but parents in Cluster 2 had a lower percentage of receiving limited education.

**Table 1:** Characteristics of all included twins overall and in the two clusters. The *p-values* are for differences between Clusters 1 and 2 by Chi-squared test or univariable linear regression accounting for twin sampling.

| Characteristic | n (%) / mean ± SD | | | *p-value* |
| --- | --- | --- | --- | --- |
|  | Overall<br>(individual twin n=1804) | Cluster 1<br>(individual twin n=736) | Cluster 2<br>(individual twin n=1068) |  |
| **Sex** |  |  |  | 0.16 |
| Male | 745 (41.3) | 289 (39.3) | 456 (42.7) |  |
| Female | 1059 (58.7) | 447 (60.7) | 612 (57.3) |  |
| **Zygosity** |  |  |  | 0.92 |
| Monozygotic | 615 (34.1) | 252 (34.2) | 363 (34.0) |  |
| Dizygotic | 1105 (61.3) | 448 (60.9) | 657 (61.5) |  |
| Unknown | 84 (4.7) | 36 (4.9) | 48 (4.5) |  |
| **Smoking** |  |  |  | 0.03 |
| Never | 994 (55.1) | 405 (55) | 589 (55.2) |  |
| Former | 191 (10.6) | 78 (10.6) | 113 (10.6) |  |
| Occasional | 205 (11.4) | 66 (9.0) | 139 (13.0) |  |
| Current | 414 (23.0) | 187 (25.4) | 227 (21.3) |  |
| **Work** |  |  |  | <0.0001 |
| Full-time work | 880 (48.8) | 409 (55.6) | 471 (44.1) |  |
| Part-time work | 280 (15.5) | 94 (12.8) | 186 (17.4) |  |
| Irregular work | 239 (13.3) | 76 (10.3) | 163 (15.3) |  |
| Not working | 405 (22.5) | 157 (21.3) | 248 (23.2) |  |
| **Secondary level school** |  |  |  | <0.0001 |
| Vocational | 486 (26.9) | 262 (35.6) | 224 (21.0) |  |
| Senior high school | 1222 (67.7) | 439 (59.7) | 783 (73.3) |  |
| None | 96 (5.3) | 35 (4.8) | 61 (5.7) |  |
| **Parental education** |  |  |  | <0.0001 |
| Limited | 922 (51.1) | 429 (58.3) | 493 (46.2) |  |
| Intermediate | 410 (22.7) | 155 (21.1) | 255 (23.9) |  |
| High | 472 (26.2) | 152 (20.7) | 320 (30.0) |  |
| **Age** | 24.07 (1.7) | 24.15 (1.7) | 24.01 (1.7) | 0.10 |
| **GBI in young adulthood** | 4.42 (4.7) | 4.05 (4.4) | 4.67 (4.8) | 0.01 |

### Linear elastic net regression model

After full adjustment (Table 2), within the sample of all twins, six land use exposures: low-density residential land use in a 100 m buffer, natural land use in a 100 m buffer, high-density residential land use in a 300 m buffer, infrastructures land use in a 300 m buffer, natural land use in a 300 m buffer, and high-density residential land use in a 500 m buffer were significant enough to be captured by the linear elastic net regression model in assessing their relationship with GBI. The number of selected land use exposures increased to 11 in Cluster 1 model (suburban), while surprisingly there were no land use exposures remaining in Cluster 2 (city center) model. The pattern of coefficients including the effect size and direction was relatively heterogeneous. The coefficients for low-density residential land use in a 100 m buffer were the same (coefficient: -0.011) between the overall and Cluster 1 models. Additionally, infrastructure land use in a 300 m buffer and high-density residential land use in a 500 m buffer were captured by both the overall and Cluster 1 models, but the effect size or direction are quite heterogeneous. Agricultural residential land use in a 100 m buffer contributed to GBI to the largest degree in Cluster 1 model (coefficient: 0.097). The GBI was linearly correlated with none land use exposures in Cluster 2.

**Table 2:**
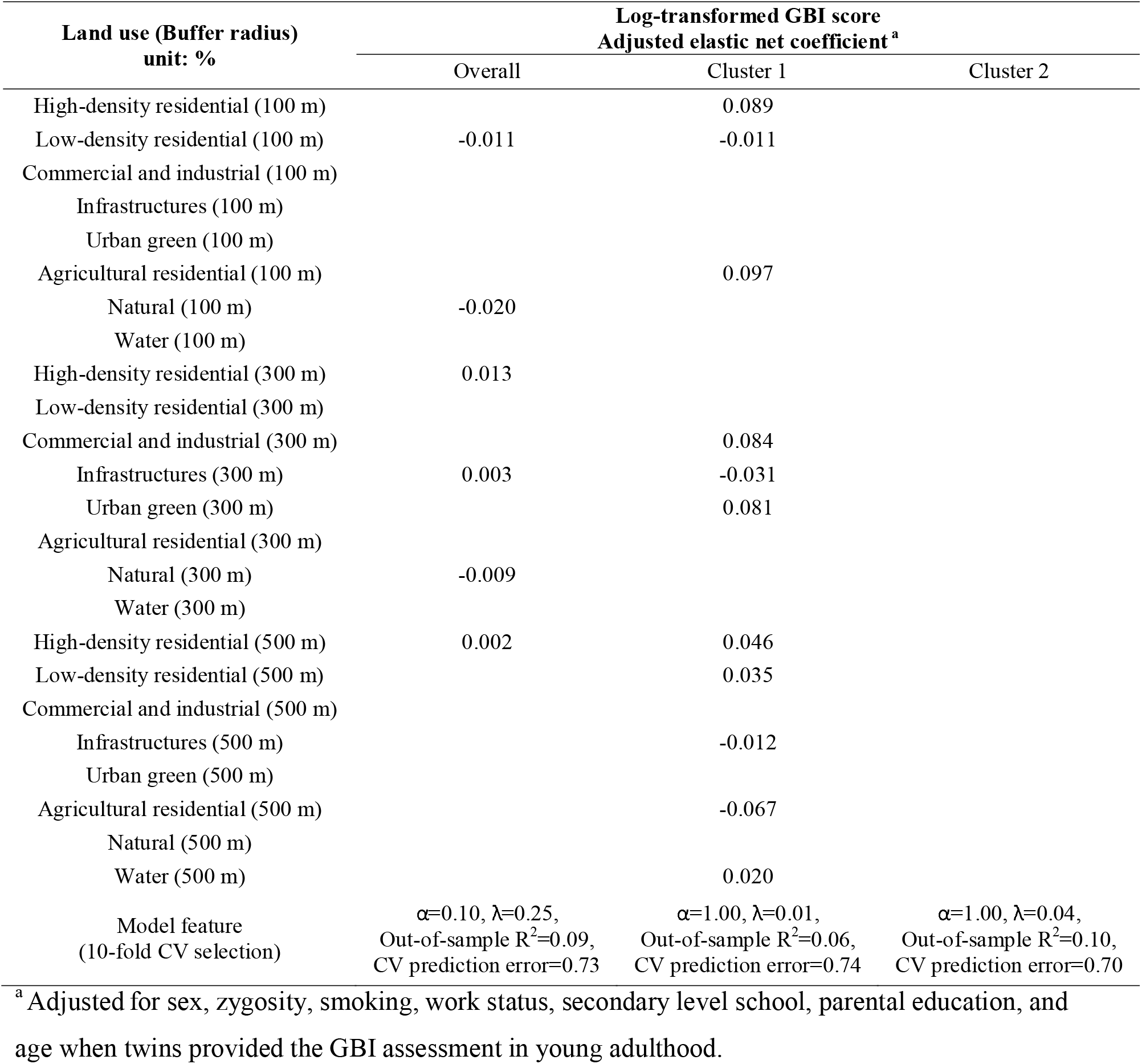
Multiple-exposure elastic net penalized regression for associations between land use and GBI. The remaining coefficients were significant enough to be selected.

| Land use (Buffer radius)<br>unit: % | Log-transformed GBI score<br>Adjusted elastic net coefficient <sup>a</sup> |  |  |
| --- | --- | --- | --- |
|  | Overall | Cluster 1 | Cluster 2 |
| High-density residential (100 m) |  | 0.089 |  |
| Low-density residential (100 m) | -0.011 | -0.011 |  |
| Commercial and industrial (100 m) |  |  |  |
| Infrastructures (100 m) |  |  |  |
| Urban green (100 m) |  |  |  |
| Agricultural residential (100 m) |  | 0.097 |  |
| Natural (100 m) | -0.020 |  |  |
| Water (100 m) |  |  |  |
| High-density residential (300 m) | 0.013 |  |  |
| Low-density residential (300 m) |  |  |  |
| Commercial and industrial (300 m) |  | 0.084 |  |
| Infrastructures (300 m) | 0.003 | -0.031 |  |
| Urban green (300 m) |  | 0.081 |  |
| Agricultural residential (300 m) |  |  |  |
| Natural (300 m) | -0.009 |  |  |
| Water (300 m) |  |  |  |
| High-density residential (500 m) | 0.002 | 0.046 |  |
| Low-density residential (500 m) |  | 0.035 |  |
| Commercial and industrial (500 m) |  |  |  |
| Infrastructures (500 m) |  | -0.012 |  |
| Urban green (500 m) |  |  |  |
| Agricultural residential (500 m) |  | -0.067 |  |
| Natural (500 m) |  |  |  |
| Water (500 m) |  | 0.020 |  |
| Model feature<br>(10-fold CV selection) | $\alpha=0.10, \lambda=0.25,$<br>Out-of-sample $R^2=0.09,$<br>CV prediction error=0.73 | $\alpha=1.00, \lambda=0.01,$<br>Out-of-sample $R^2=0.06,$<br>CV prediction error=0.74 | $\alpha=1.00, \lambda=0.04,$<br>Out-of-sample $R^2=0.10,$<br>CV prediction error=0.70 |

<sup>a</sup> Adjusted for sex, zygosity, smoking, work status, secondary level school, parental education, and age when twins provided the GBI assessment in young adulthood.

### XGBoost model

We listed the top 10 most important factors with SHAP values in each XGBoost model (Figure 2). For example, the top 10 in the overall models are natural land use in a 100 m buffer, commercial and industrial land use in a 300 m buffer, low-density residential land use in a 300 m buffer, low-density residential land use in a 500 m buffer, natural land use in a 500 m buffer, high-density residential land use in a 500 m buffer, urban green land use in a 500 m buffer, and commercial and industrial land use in a 500 m buffer (in order). Covariates were not listed and are not shown in the figure. The curve of SHAP values suggested non-linear attribution of each land use exposure on GBI. Industrial and commercial use in a 300 m buffer and low-density residential land use in a 500 m buffer were the most important land use exposures in Cluster 1 (suburban) and Cluster 2 (city center) models, respectively.

**Figure 2:**
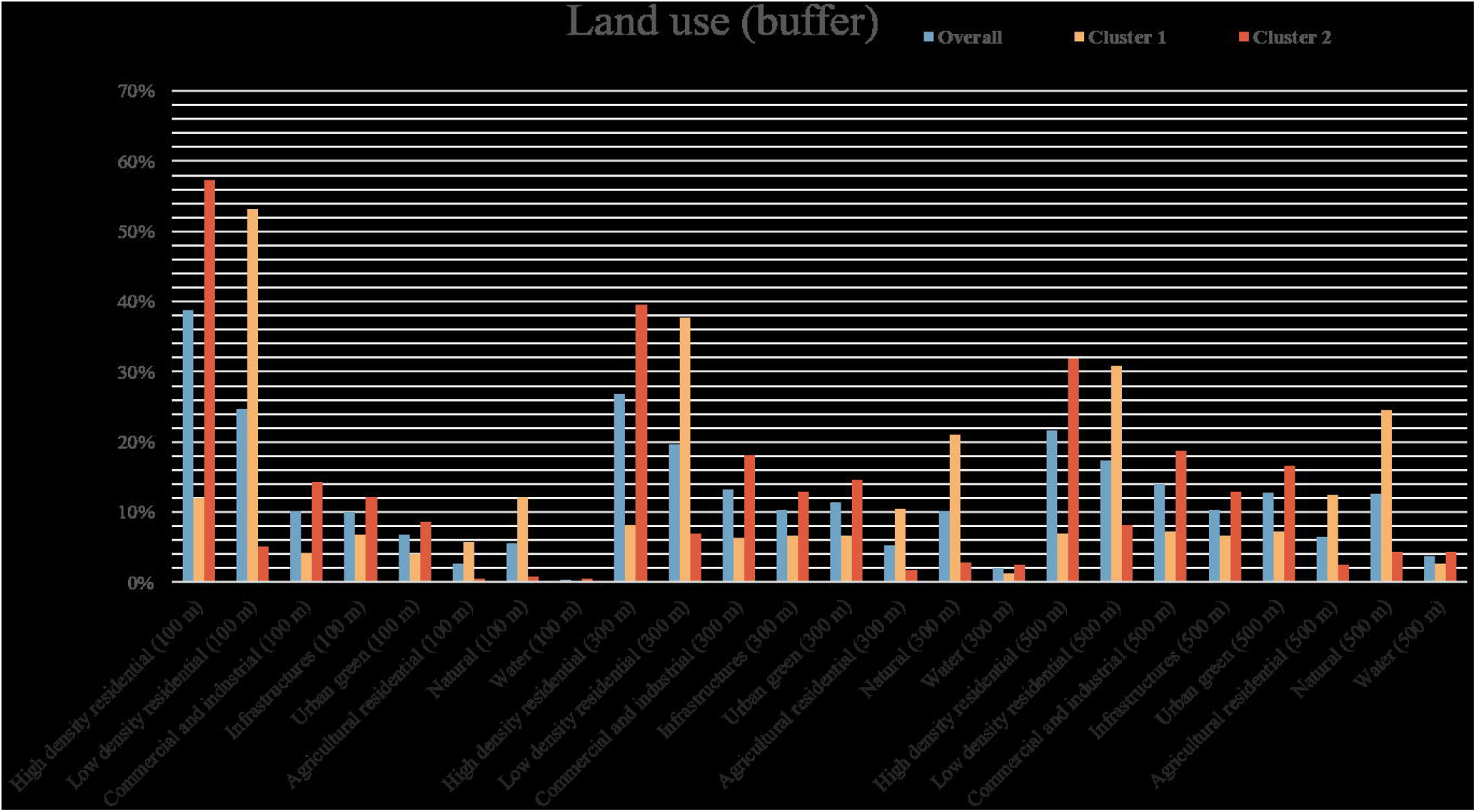
Shapley (SHAP) plots illustration of the top 10 most influential exposures in the overall (A), Cluster 1 (B), and Cluster 2 (C) XGBoost models. Covariates were included in the models but suppressed in plots to highlight land use exposures.

For nature land use in a 100 m buffer in the overall model, there was an obvious decline of SHAP value between 0 and ∼10%. Then, the value increased when its percentage passed ∼10% and, after the percentage was greater than ∼22%, the curve was relatively flat. A similar pattern was also observed in the plot of industrial and commercial land use in a 300 m buffer in Cluster 1 model. However, the curve of low-density residential land use in a 500 m buffer was always relatively flat in Cluster 2 model.

### Model performance and comparison

The standard deviations (SD) of the log-transformed GBI score were 0.8825, 0.8851, and 0.8774 among the overall, Cluster 1’s and Cluster 2’s twins. The training and testing RMSE are shown in Table 3, there are no major differences between the two types of models and clusters, and they are mostly lower than the SDs of the log-transformed GBI score, implying good model performance.

**Table 3:** Model performance via root-mean-squared error (RMSE) for linear elastic net penalized regression and XGBoost models

| Model accuracy |  | Training RMSE | Testing RMSE |
| --- | --- | --- | --- |
| Linear elastic net<br>penalized regression | Overall | 0.840 | 0.817 |
|  | Cluster 1 | 0.825 | 0.817 |
|  | Cluster 2 | 0.835 | 0.782 |
| XGBoost | Overall | 0.833 | 0.891 |
|  | Cluster 1 | 0.734 | 0.879 |
|  | Cluster 2 | 0.804 | 0.891 |

### Post-hoc linear regression

The results of linear regression in the overall and the two separated cluster models are presented in Supplemental Table 3. In crude Cluster 1 (suburban) model, a higher land use mix index within a 300 m buffer was significantly associated with higher log-transformed GBI scores (beta: 0.51, 95% CI: 0.02, 1.01). After adjustment, there was no significant association.

## Discussion

Based on 1804 twins from the FinnTwin12 study with information on residential geocodes linked to land use characteristics, we identified two clusters with notable differences in the percentage of high-density residential, low-density residential, and natural land use. By two types of models, both linear and non-linear relationships between land use and depressive symptoms were discovered to exist. In the linear elastic net penalized regression model among overall twins and Cluster 1 (suburban)’s twins, there was a heterogeneous pattern in selected subsets, effect sizes, and effect directions. In the Cluster 1 model, the agricultural residential land use in a 100 m buffer was associated with depressive symptoms with the largest relative effect size. In contrast, no land use exposures were significant enough to be attributed to depressive symptoms in Cluster 2, which was typical of city or town centers. Between the overall, Cluster 1, and Cluster 2 XGBoost models, the ranks of land use exposures’ importance on depressive symptoms were also heterogeneous and the most important were natural land use in a 100 m buffer, commercial and industrial land use in a 300 m buffer, and low-density residential in a 500 buffer, respectively. As a hypothesis-generating study from the Equal-life project, elements such as population heterogeneity, environmental interaction, and characteristics of the effect (such as linearity) should be more considered in future analyses between land use, as well as the broad urban exposome, and depressive symptoms.

First, the clustering analysis revealed a specific pattern in urbanization, and twins from Clusters 1 and 2 mostly lived in the “suburbs” and “city or town centers”, respectively. The land use exposures appear to less important to depressive symptoms among people living in the city or town centers. The possible mechanisms may be through differential healthcare service access, social needs, transportation connectedness, or neighborhood environment.^14,40,41^ For example, living in the suburbs usually requires longer house-to-job distances, which has been found to be associated with poorer mental health.^40^ The longer job commutes implied more need for transportation facilities, and, similar to our linear elastic net regression model, the higher percentage of infrastructure land use was related to less depressive symptoms in Cluster 1 (suburban). Nevertheless, Pelgrims et al. detected no significant association, after fully adjustment, between green surrounding, street corridor and canyon effect, and depressive disorder among participants living in the highly urbanized Brussels, Belgium.^42^ We did not intend to distinguish people with an arbitrary binary classification, instead, we promote the hypothesis that the relationship between land use and depressive symptoms exists in the specific land use context.

More broadly, many land use exposures, that signaled urbanization, were either selected by the penalized model or ranked in the top 10 most important in XGBoost, suggesting its effect on depressive symptoms. A 2020 review found the protective effect of urbanization on depression in three Chinese studies, while four other countries’ studies had opposite findings due to different geographic regions and income levels.^43^ An increasing trend in depression prevalence among young adults and those who lived in rural areas with low population density was observed in a longitudinal Germany nationwide survey.^44^ However, Morozov indicated that urbanization adversely affected mental health via several factors including noise and visual aggressiveness of the environment in Russia.^45^ Our conventional analysis with the land use mix index indicated null results, and previous literature also shows inconsistent findings^13–15^, which increases the interest in deeper assessment. There may be conjunct or nonadditive relationships within land use or broad urban living environments. For instance, the urban heat island, with a higher regional temperature in urban areas than in surrounding rural areas, has been shown to be differentially influenced by many land use factors, in which expansion of built-up area increased but water areas reduced the regional temperature^46^, and moreover the urban heat island increases the risk of depression.^43^

Including multiple land use exposures in a single analysis platform allows us to disentangle the individual effects and assess the complex relationships. The linear elastic net penalized regression models selected a subset of the most influential land use exposures, exerted combined effects, and avoided the risk of multicollinearity and overfitting.^47^ Because we aim to reveal the relationship instead of prediction, we did not refill the land use exposures to the normal regression model and the interpretation of effect size was weakened. Lenters et al. have applied this approach to prenatal chemical exposures to solve the interconnected effects of mixtures.^31^ We also observed the nonlinear relationship via the interpretable SHAP visualization from XGBoost, but, like Ohanyan and colleagues’ studies, we did not straightforwardly assess the interaction due to modest effect sizes and other factors.^18,48^ Previous applications of this machine learning method improved the prediction of air quality and enhanced the forecast of air quality in China.^34,49^ Ma et al. also compared the prediction accuracy between XGBoost and Lasso penalized regression models^49^, while, in our study, we wished to observe the intricate effects instead of comparing accuracy, so we used RMSE, not AUC, to evaluate model performance. Another Chinese study also explored the nonlinear effect between the built and social environments and bus use among the older adults.^35^ The utility of multiple machine learning algorithms provides a preliminary sketch of the labyrinthine relationship between urban land use and depression symptoms.

Clustering analysis focused on multiple land use exposures and facilitates the segmentation of residents for tailored epidemiological assessment of the effect of land use on depressive symptoms and customizes further improvement and intervention. The differential pattern of urban land use environment was very obvious in our findings. Methodologically, clustering analysis has gained increasing attention in the field of exposure science. Tognola and colleagues clustered children in France by exposure to extremely low-frequency magnetic fields^50^, and another study developed a novel workflow in clustering with multiple features including specific and general external exposomes and identified sub-populations in type-2 diabetes patients.^51^ Moreover, wildlife necropsy data has also been clustered for syndromic surveillance of any new zoonotic outbreak^52^, which engaged the health interaction between humans and animals and is an example of the application of this method in “One Earth”.

There are some limitations in our studies. First, the information on depression symptoms was obtained before 2012, so the potential causality and direction are unable to be confirmed due to temporality. Second, compared to previous similar studies, the sample size is relatively small. Although the two machine learning methods are able to shrink the overfitting due to the small sample size, we still need to be cautious about the findings. This study is a pilot study for exploration, and further follow-up studies are welcome to strengthen the evidence.

## Conclusion

This study is the first, to our knowledge, to investigate the complex relationship between multiple urban land use exposures and depressive symptoms in young adulthood. The pluralistic multi-model inferences selected or prioritized the more important urban land use exposures to depressive symptoms and revealed the linear and nonlinear relationships, which advances the conventional assessment with a single index. Clustering analysis showed a notably heterogeneous pattern in this relationship between participants with different land use environments, implying the effects are under a specific context. Due to sample size, model characteristics, and temporality, our finding interpretation is cautious at present, and more efforts are warranted to corroborate.

## Data Availability

The FinnTwin12 data is not publicly available due to the restrictions of informed consent. However, the FinnTwin12 data is available through the Institute for Molecular Medicine Finland (FIMM) Data Access Committee (DAC) for authorized researchers who have IRB/ethics approval and an institutionally approved study plan. To ensure the protection of privacy and compliance with national data protection legislation, a data use/transfer agreement is needed, the content and specific clauses of which will depend on the nature of the requested data.

## Acknowledgements

We would like to appreciate the analysis support from Gabin Drouard (University of Helsinki) and Tianze Lin (Southern University of Science and Technology). We would like to Dr. Maria Foraster from the Barcelona Institute for Global Health (ISGlobal) for her contribution to data acquisition on land use. FinnTwin12 wishes to thank all participating twins, their parents, and teachers.

## Ethical statement

The ethics committee of the Department of Public Health of the University of Helsinki (Helsinki, Finland), the ethics committee of the Helsinki University Central Hospital District (Helsinki, Finland), and the Institutional Review Board of Indiana University (Bloomington, Indiana, USA) approved the FinnTwin12 study protocol. All participants and their parents/legal guardians gave informed written consent to participate in the study. The authors assert that all procedures contributing to this work comply with the ethical standards of the relevant national and institutional committees on human experimentation and with the Helsinki Declaration of 1975, as revised in 2008.

