## Supplemental Table 1-3 and Figure 1-4 for "Exposome approaches to assessing the association between urban land use environment and depressive symptoms in young adulthood: a FinnTwin12 cohort study"

Zhiyang Wang et al. – Online supplemental material

Content of Supplemental Material

Supplemental Table 1: Characteristics of training and testing subsets

Supplemental Table 2: Land use characteristics of overall twins and in two clusters

Supplemental Table 3: Linear regression between land use mix index and GBI in young adulthood

Supplemental Figure 1: 2D scatter plot of K-means clustering

Supplemental Figure 2: Distribution of twins in greater Helsinki areas in 2012 by Clusters 1 (red) and 2 (green)

Supplemental Figure 3: Matrix of Pearson correlations between land use exposures based on the training subsets. The number indicates the correlation coefficient, the size of the circle indicates the size of the coefficient, and the blank (no color) indicates the lack of significance of the correlation coefficient.

Supplemental Table 1: Characteristics of training and testing subsets

| **Characteristic** | **n (%) / mean ± SE** | |
| --- | --- | --- |
|  | Training  (individual twin n=1215) | Testing (individual twin n=589) |
| **Sex** |  |  |
| Male | 508 (41.8) | 237 (40.2) |
| Female | 707 (58.2) | 352 (59.8) |
| **Zygosity** |  |  |
| Monozygotic | 392 (32.3) | 223 (37.9) |
| Dizygotic | 763 (62.8) | 342 (58.1) |
| Unknown | 60 (4.9) | 24 (4.1) |
| **Smoking** |  |  |
| Never | 654 (53.8) | 340 (57.7) |
| Former | 137 (11.3) | 54 (9.2) |
| Occasional | 149 (12.3) | 56 (9.5) |
| Current | 275 (22.6) | 139 (23.6) |
| **Work** |  |  |
| Full-time work | 606 (49.9) | 274 (46.5) |
| Part-time work | 166 (13.7) | 114 (19.4) |
| Irregular work | 159 (13.1) | 80 (13.6) |
| Not working | 284 (23.4) | 121 (20.5) |
| **Secondary level school** |  |  |
| Vocational | 332 (27.3) | 154 (26.2) |
| Senior high school | 821 (67.6) | 401 (68.1) |
| None | 62 (5.1) | 34 (5.8) |
| **Parental education** |  |  |
| Limited | 644 (53.0) | 278 (47.2) |
| Intermediate | 271 (22.3) | 139 (23.6) |
| High | 300 (24.7) | 172 (29.2) |
| **Age** | 24.14 (1.7) | 23.92 (1.7) |
| **GBI in young adulthood** | 4.49 (4.7) | 4.28 (4.6) |

Supplemental Table 2: Land use characteristics of overall twins and in the two clusters

| Land use (Buffer) unit: % | Variable name | mean ± SD | | | Ratio of means  between two clusters ^a^ |
| --- | --- | --- | --- | --- | --- |
|  |  | Overall  (individual twin n=1804) | Cluster 1  (individual twin n=736) | Cluster 2  (individual twin n=1068) |  |
| High-density residential (100 m) | urbanhigh_100_2012 | 0.388 (0.330) | 0.120 (0.217) | 0.573 (0.261) | 4.78 |
| Low-density residential (100 m) | urbanlow_100_2012 | 0.247 (0.324) | 0.532 (0.311) | 0.051 (0.130) | 10.43 |
| Commercial and industrial (100 m) | com_ind_100_2012 | 0.102 (0.192) | 0.042 (0.113) | 0.143 (0.222) | 3.40 |
| Infrastructures (100 m) | infrast_100_2012 | 0.100 (0.078) | 0.068 (0.044) | 0.122 (0.089) | 1.79 |
| Urban green (100 m) | green_urb_100_2012 | 0.068 (0.116) | 0.040 (0.089) | 0.087 (0.129) | 2.18 |
| Agricultural residential (100 m) | agric_100_2012 | 0.027 (0.094) | 0.058 (0.136) | 0.005 (0.033) | 11.60 |
| Natural (100 m) | natural_100_2012 | 0.055 (0.133) | 0.121 (0.181) | 0.009 (0.045) | 13.44 |
| Water (100 m) | water_100_2012 | 0.004 (0.028) | 0.001 (0.010) | 0.006 (0.036) | 6.00 |
| High-density residential (300 m) | urbanhigh_300_2012 | 0.268 (0.217) | 0.082 (0.103) | 0.396 (0.178) | 4.83 |
| Low-density residential (300 m) | urbanlow_300_2012 | 0.196 (0.214) | 0.377 (0.202) | 0.070 (0.103) | 5.39 |
| Commercial and industrial (300 m) | com_ind_300_2012 | 0.133 (0.141) | 0.064 (0.090) | 0.181 (0.150) | 2.83 |
| Infrastructures (300 m) | infrast_300_2012 | 0.103 (0.066) | 0.066 (0.037) | 0.129 (0.069) | 1.95 |
| Urban green (300 m) | green_urb_300_2012 | 0.114 (0.114) | 0.067 (0.086) | 0.146 (0.119) | 2.18 |
| Agricultural residential (300 m) | agric_300_2012 | 0.053 (0.120) | 0.105 (0.164) | 0.018 (0.053) | 5.83 |
| Natural (300 m) | natural_300_2012 | 0.102 (0.163) | 0.211 (0.197) | 0.028 (0.068) | 7.54 |
| Water (300 m) | water_300_2012 | 0.020 (0.062) | 0.013 (0.047) | 0.025 (0.070) | 1.92 |
| High-density residential (500 m) | urbanhigh_500_2012 | 0.217 (0.173) | 0.070 (0.077) | 0.319 (0.146) | 4.56 |
| Low-density residential (500 m) | urbanlow_500_2012 | 0.174 (0.171) | 0.308 (0.165) | 0.082 (0.098) | 3.76 |
| Commercial and industrial (500 m) | com_ind_500_2012 | 0.140 (0.121) | 0.072 (0.081) | 0.187 (0.121) | 2.60 |
| Infrastructures (500 m) | infrast_500_2012 | 0.103 (0.063) | 0.066 (0.040) | 0.129 (0.064) | 1.95 |
| Urban green (500 m) | green_urb_500_2012 | 0.128 (0.108) | 0.072 (0.084) | 0.166 (0.106) | 2.31 |
| Agricultural residential (500 m) | agric_500_2012 | 0.065 (0.123) | 0.124 (0.162) | 0.025 (0.059) | 4.96 |
| Natural (500 m) | natural_500_2012 | 0.126 (0.169) | 0.246 (0.190) | 0.043 (0.081) | 5.72 |
| Water (500 m) | water_500_2012 | 0.037 (0.081) | 0.027 (0.068) | 0.043 (0.089) | 1.59 |

^a^ The larger of the means is used in the numerator in the ratio**.**

Supplemental Table 3: Linear regression between land use mix index and GBI in young adulthood

| Land use mix index | mean ± SD | Log-transformed GBI scores in young adulthood | |
| --- | --- | --- | --- |
|  |  | Unadjusted beta (95% CI) | Adjusted beta (95% CI) ^a^ |
| *Overall (individual twin n=1804)* | | | |
| Within 100 m buffer | 0.38 (0.14) | 0.11 (-0.17, 0.39) | 0.17 (-0.10, 0.44) |
| Within 300 m buffer | 0.60 (0.12) | 0.17 (-0.18, 0.52) | 0.14 (-0.19, 0.48) |
| Within 500 m buffer | 0.67 (0.11) | 0.30 (-0.08, 0.68) | 0.25 (-0.11, 0.62) |
| *Cluster 1 (individual twin n=736)* | | | |
| Within 100 m buffer | 0.38 (0.15) | 0.02 (-0.39, 0.44) | 0.12 (-0.28, 0.52) |
| Within 300 m buffer | 0.59 (0.13) | 0.51 (0.02, 1.01)* | 0.46 (-0.01, 0.92) |
| Within 500 m buffer | 0.66 (0.13) | 0.50 (-0.00, 1.01) | 0.42 (-0.07, 0.91) |
| *Cluster 2 (individual twin n=1068)* | | | |
| Within 100 m buffer | 0.38 (0.14) | 0.16 (-0.21, 0.53) | 0.18 (-0.18, 0.54) |
| Within 300 m buffer | 0.60 (0.11) | -0.25 (-0.73, 0.23) | -0.27 (-0.73. 0.19) |
| Within 500 m buffer | 0.68 (0.10) | -0.08 (-0.63, 0.48) | -0.14 (-0.67, 0.40) |

^a^ Adjusted for sex, zygosity, smoking, work status, secondary level school, parental education, and age when twins provided the GBI assessment in young adulthood.

* *p-value* <0.05

Supplemental Figure 1: 2D scatter plot of K-means clustering

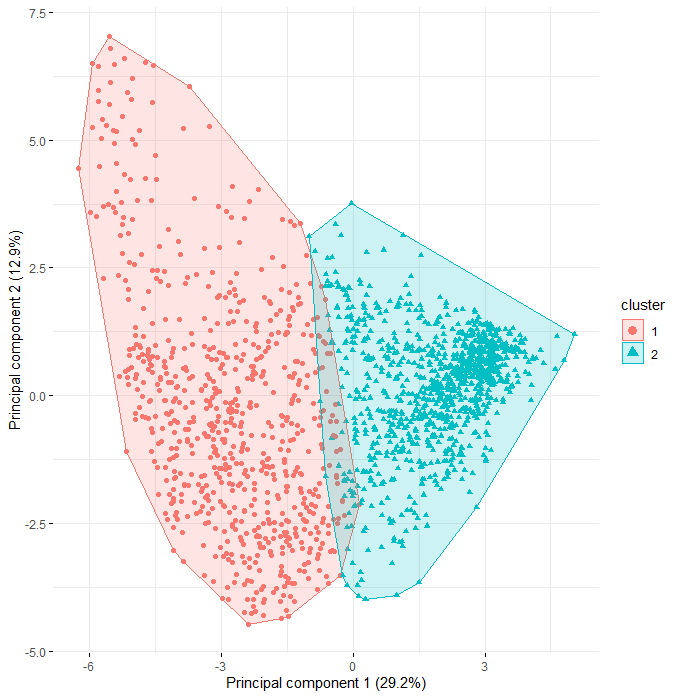

Supplemental Figure 2: Distribution of twins in greater Helsinki areas in 2012 by Clusters 1 (red) and 2 (green)

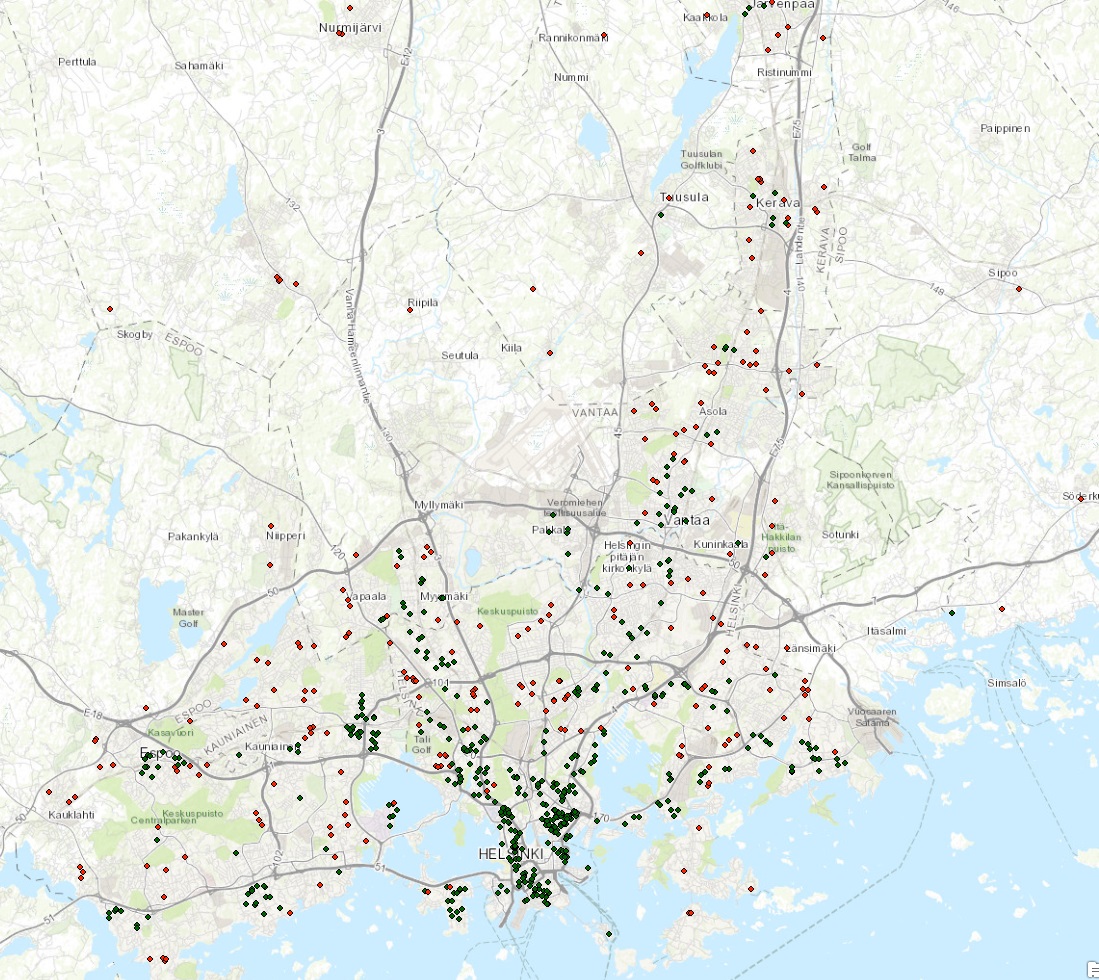

Supplemental Figure 3: Matrix of Pearson correlations between land use exposures based on the training subsets. The number indicates the correlation coefficient, the size of the circle indicates the size of the coefficient, and the blank (no color) indicates the lack of significance of the correlation coefficient.

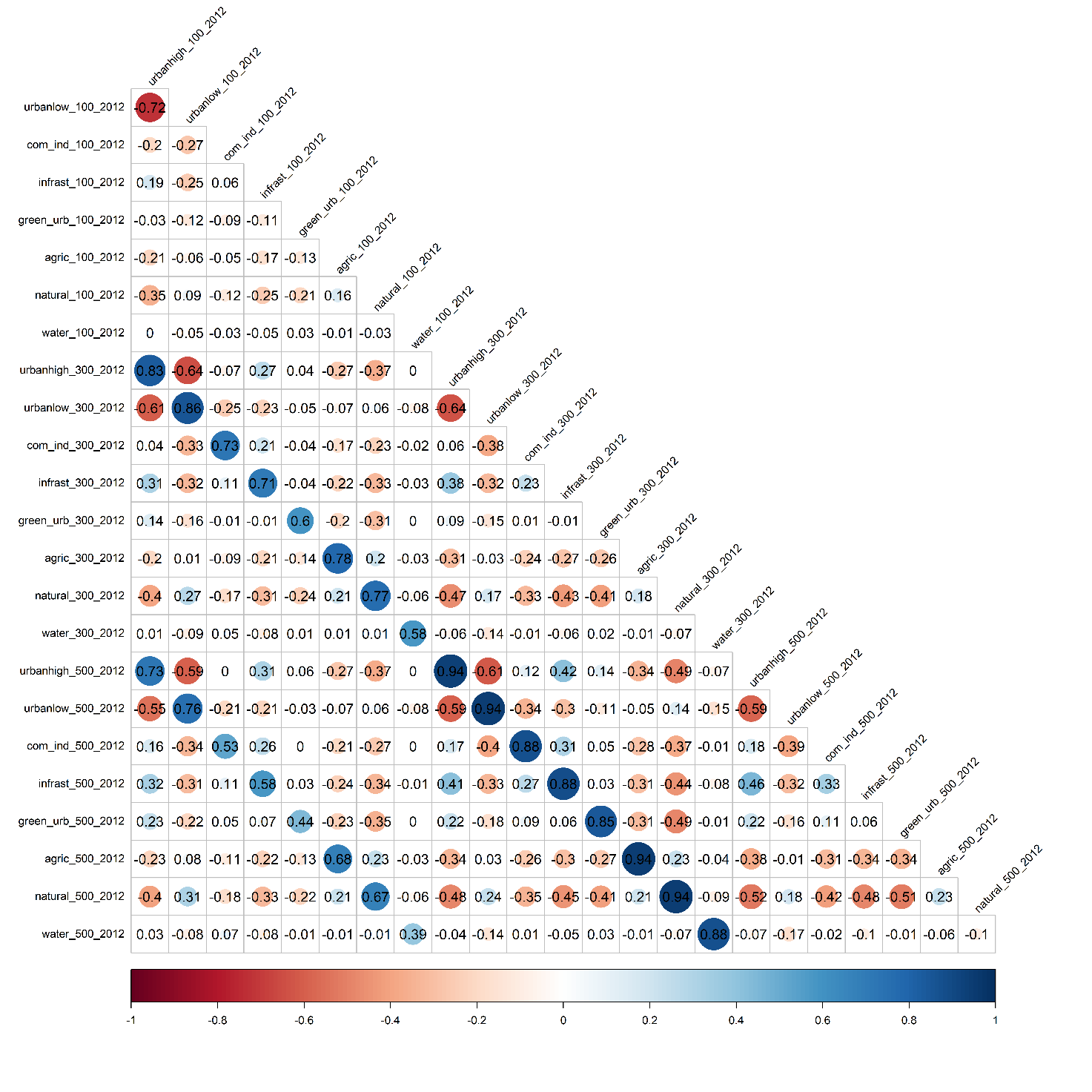
